# Stakeholder and end-user perspectives to improve the Context-sensitive Positive Health Questionnaire (CPHQ) to measure broad health

**DOI:** 10.1101/2025.08.01.25332588

**Authors:** Mirte Boelens, Cheryl Roumen, Esther J. Bloemen-van Gurp, John Dierx, Lenny Nahar-van Venrooij, Tim van Zutphen, Pien M.R. Christiaanse, Jaron Schnitzer, M. Elske van den Akker-van Marle, Miriam J.J. de Kleijn, Marja van Vliet, Jessica C. Kiefte-de Jong, Marieke D. Spreeuwenberg

**Affiliations:** Leiden University Medical Center/Health Campus The Hague, Department of Public Health and Primary Care, The Hague, The Netherlands; Department of Health Services Research, Care and Public Health Research Institute (CAPHRI), Faculty of Health Medicine and Life Sciences, Maastricht University, Maastricht, The Netherlands; Zuyd University of Applied Sciences, Research Center of community care, Heerlen, The Netherlands; Fontys University of Applied Sciences, Center of Expertise Health, Eindhoven, The Netherlands; Avans University of Applied Science, Centre of Expertise Perspective in Health, Research Group Equal Chances on Healthy Choices, Breda, Netherlands; Jeroen Bosch Academy Research, Jeroen Bosch Hospital, ’s-Hertogenbosch, Netherlands; Tranzo Scientific Centre for Care and Wellbeing, Tilburg University, Tilburg, the Netherlands; Department of Sustainable Health, Faculty Campus Fryslân, University of Groningen, Leeuwarden, the Netherlands; Department of Biomedical Data Sciences, Section of Medical Decision-Making, Leiden University Medical Center, Leiden, The Netherlands; Institute for Positive Health, Utrecht, The Netherlands

**Keywords:** Health status measurement, broad health, Positive Health, Capability Approach, Context-Sensitive Positive Health Questionnaire

## Abstract

**Introduction:** Broad health concepts are increasingly embedded in healthcare and other domains. Therefore, in a previous study, we developed and validated the Context-Sensitive Positive Health Questionnaire (CPHQ). The CPHQ is based on Positive Health, the Capability Approach and on perspectives of stakeholders and end-users, including those with a lower socioeconomic position (SEP) with the aim to validly measure broad health. Stakeholders and end-users were involved in the development process. However pronfessionals working in healthcare, welfare or policy domain were not sufficiently involved while their perspective on how to measure broad health is needed to support broader applicability and acceptability.

**Objective:** We aimed to refine the 32-item CPHQ by incorporating the perspectives of a broader group of stakeholders across healthcare, welfare and policy domains, researchers, as well as patients and citizens, including those from a lower SEP, so that the instrument is suitable for use in diverse domains and population groups.

**Methods:** We conducted nine focus group discussions with stakeholders from healthcare, welfare, and policy domains, research, patients and (lower SEP) citizens. An adapted COMET method analysis and qualitative analysis were used followed by a member check took to validate the changes.

**Results:** The refined CPHQ consists of 28 items. Items were omitted, merged or added. Participants emphasized the importance of avoiding normative phrasing and including subjectively interpretable items. Items reflecting aspects of experienced health (e.g., “I feel happy” or “I feel healthy”) were prioritized, highlighting the relevance of functionings. At the same time, views diverged on the inclusion and wording of items related to exclusion, political representation and sense of belonging.

**Conclusion:** The CPHQ refinement involved a wide range of stakeholders including end-users. This led to a 28-item CPHQ to measure broad health. Future research in different populations and settings is needed to validate and possibly shorten the refined CPHQ.

## Introduction

There is growing recognition that health encompasses more than the absence of disease. In response to critiques of the World Healthcare Organization’s (WHO) definition of health, which is the complete state of physical, mental and social well-being several approaches to measure broad health have been proposed (World Health Organization). In 2011, Huber proposed a more dynamic conceptualization of health as “the ability to adapt and to self-manage in the face of life’s physical, emotional, and social challenges” (Huber, Knottnerus et al. 2011). This was operationalized into the concept of Positive Health, which reflects health as a broad construct comprising six dimensions: Bodily functions, Mental well-being, Meaningfulness, Quality of life, Participation, Daily functioning. Positive Health is about what people themselves perceive to constitute their health, and what gives meaning to their lives to enable self-management and resilience (Huber, van Vliet et al. 2016). The Capability Approach, as developed by Sen and Nussbaum, also offers a dynamic and context-sensitive framework by focusing on individuals’ opportunities to achieve what they value in terms of health (Sen 1993, Nussbaum 2011). The Capability approach distinguishes between actual states of health called “functionings” (e.g. feeling healthy or feeling happy) and “capabilities” that enable people to achieve these states of health. Health, in this view, is not only defined by actual outcomes, but also by the capabilities to attain them. The conversion of resources into capabilities depends on “conversion factors”. “Conversion factors” can be personal (e.g. sex or reading skills), social (e.g. public policies, social norms or discriminating practices) or environmental (e.g. climate or geographical region)(Robeyns 2005, Robeyns 2013).

Within Dutch healthcare policy, there is a growing emphasis on health as a broad conceptualization of health in which the ability to adapt is included (Huber, Knottnerus et al. 2011). Accordingly, policy documents and health programs within the healthcare and social domain have been established in line with this framework. However, so far, no adequately validated questionnaires seem to be available to evaluate the impact of these health programs and interventions (Johansen, Loorbach et al. 2018, Dutch National Health Care Institute & Dutch Health Care Authority 2020, Ministry of Health Welfare and Sports 2020, Zonneveld, Glimmerveen et al. 2022).

Therefore, attempts have been made to develop a valid questionnaire to measure broad health. A previous attempt showed that the 42-item My Positive Health questionnaire, originally developed as a dialogue tool (https://www.iph.nl/en/positive-health/conversation-tools/), was not suitable as a validated measurement instrument (Prinsen and Terwee 2019, Van Vliet, Doornenbal et al. 2021). Subsequently, this questionnaire was reordered and reduced to a 17-item questionnaire using factor analysis (Van Vliet, Doornenbal et al. 2021). The 17-item questionnaire showed adequate psychometric properties(Van Vliet, Doornenbal et al. 2021). However, in line with current policy and intervention approaches, and as suggested by the International Union for Health Promotion and Education (IUHPE), it would be more valuable to develop a measurement instrument based on Positive Health, supplemented with contextual factors such as political, environmental, and social aspects (Shilton, Sparks et al. 2011). Additionally, a measurement instrument developed in this way is expected to align well with current Dutch governmental policies and intervention approaches within the healthcare and welfare and policy settings. As a result, it is likely to be both sensitive and responsive in capturing the impact of health interventions.

Therefore, in a previous study, we developed and validated a context-sensitive Positive Health questionnaire (CPHQ) as a measurement instrument based on the principles of Positive Health and grounded in the Capability Approach by accounting for differences in capabilities and conversion factors influencing health (Doornenbal, van Zutphen et al. 2024). This CPHQ consists of 32 items and comprises 11 dimensions; relaxation, autonomy, fitness, perceived environmental safety, exclusion, social support, financial resources, political representation, health literacy, resilience, and enjoyment. This CPHQ demonstrated adequate factorial and concurrent validity. However, in the development of the CPHQ a broad range of potential users such as professionals working in the healthcare, welfare and policy sector were not sufficiently involved in the initial questionnaire development. They were consulted after focus group discussions with patients and citizens were finished. Taking into account their view on how to measure broad health is needed to support applicability and acceptance in difference sectors. Thus, to improve the applicability and acceptance of the CPHQ, further refinement based on the perspective of a more diverse group of stakeholders is needed.

The objective of this study is to further improve the CPHQ-instrument to make it more generally applicable and therefore accepted in different settings (i.e. healthcare, welfare, public health and the and policy domain) and by end-users. This is done by exploring needs and perceptions of stakeholders from healthcare, welfare, social and policy domain and by resonating with patients’ and (lower SEP) citizens’ perspectives.

## Materials and Methods

For the further development of the CPHQ measurement instrument different steps were conducted. Step 1 consisted of focus group discussions with stakeholders and end users (patients and (lower SEP) citizens). Step 2 consisted of an expert and member check after analysis of the focus group discussions. Both steps are described in more detail below. As a starting point for the current development, the CPHQ consisting of 32 items was used. This report follows the standards for reporting qualitative research as described by The Standards for Reporting Qualitative Research (SRQR) (O’Brien, Harris et al. 2014).

### Ethics

For this study, ethical approval was obtained from the Medical Ethics Review Committee of Leiden Den Haag Delft (LDD) (protocol 19035) which determined that the Medical Research Involving Human Subjects Act was not applicable. All participants gave written informed consent prior to study participation.

### Step 1: Focus group discussions

#### Preparation for focus group discussions

Before the focus group discussions were organized first a rapid literature search was conducted to identify other potential questionnaire items to add to our starting point; the CPHQ (Doornenbal et al., 2024). A search was conducted in both PubMed and Google Scholar with the search string: (“health” OR“Positive Health”) AND “measurement”. Also experts were consulted to provide suggestions for relevant questionnaires, such as the Positive Health measurement scale (Van Vliet, Doornenbal et al. 2021), Mijnkwaliteitvanleven.nl (Hendrikx, Drewes et al. 2019), The subjective wellbeing-5 dimensions (SWB-5D) questionnaire (Haspels, de Vries et al. 2023), 2023), ICEpop CAPability measure for Adults (ICECAP-A) (Al-Janabi, Flynn et al. 2012), Vita-16 (Strijk, Wendel-Vos et al. 2015), Brief Resilience Scale (BRS)(Smith, Dalen et al. 2008), Personal Wellbeing Index-Adult (PWI-A) (Khor, Fuller-Tysziewicz et al. 2020), EuroQol 5-dimensions (EQ-5D-5L) (Feng, Kohlmann et al. 2021), Multicultural Quality of Life Index (MQLI) (Mezzich, Cohen et al. 2011). All questionnaire items yielded from the literature search were provided hardcopy and online as a reference ‘hand book’ during the subsequent focus group discussions.

#### Focus groups discussion participants

Recruitment for the focus group discussions took place between 19-January-2023 and 16-August 2023. Ten focus groups (total of 76 participants) were organized at different locations within the Netherlands with different stakeholder groups: researcher and experts (n=8), national policy makers in health and welfare (n=6),regional policy makers in health and welfare (n=14), medical specialists and nurses (n=10), paramedics (n=8), a mixed group of professional stakeholders who could not participate during the other focus group discussions. (n=8), patients (two groups, n=8 in total), citizens (n=8), and citizens with a low socioeconomic position (n=8). A purposeful sampling approach was used.

#### Focus group discussion method

The 32 CPHQ items were presented in these focus groups on four posters (A0 size) (see Appendix 1). Items were subdivided over four posters according to the categories used in the original CPHQ, namely two posters with items concerning “perceived health” and two posters with items concerning “personal context”.

Firstly, participants received a short paper questionnaire including sociodemographic questions about their educational level, gender and work experience. Focus group discussion leaders helped participants to fill out the questionnaire if they had questions.

Round 1: Participants in each focus group were divided into groups of no more than five persons. Each group was accompanied by a researcher that guided the process. Each group discussed the four posters with the original CPHQ items. First, qualitative feedback was requested through memos on comprehensibility and relevancy of the item measuring broad health. In these sessions, the concept of broad health was defined and communicated to participants as: “The extent to which one is capable to adapt and to thrive given one’s physical, mental, social and contextual opportunities” (Doornenbal et al., 2024).

Round 2: Individual quantitative feedback was requested on the importance of the question in measuring broad health through colored stickers. The colored stickers were: green = retain question (score 1), yellow = adjust question (score 2) and orange = omit question (score 3). Each participant received 32 colored stickers of each color (1 for each CPHQ question) and assigned these individually to each item on the four posters. Each participant also received a golden ticket that could be used to mark the most important item for them.

During the focus groups, participants could consult the ‘handbook’ with items from the questionnaires that resulted from the rapid literature search to search for items that they felt were missing. Participants could also add their own items on the memo’s. After both rounds, the focus group discussion was closed with a short discussion about the most striking findings and some additional discussion points related to the preferred number of items and answer method of a measurement instrument.

Focus group discussions took between 90-120 minutes and were audio recorded.

#### Analysis of the focus group discussions

Descriptive analyses were performed for educational level, gender and work experience using IBM SPSS statistics for Windows, version 29.0 (International Business Machines Corporation, Armonk, New York).

Focus group discussions were transcribed ad verbatim and analyzed by two researchers (CR and MB). Qualitative feedback on the memos and the transcripts were analyzed on the following themes: formulation, overlap, redundancy, importance, new suggestions, duration and length of the questionnaire. Analysis of the quantitative feedback on memos for decision was done using an adapted consensus method based on the COMET handbook (Williamson et al., 2017).

For each focus group all items were scored quantitatively by counting the colored stickers scored as 1 for green, 2 for yellow and 3 for orange. Rules were defined and applied to each focus group. An item was retained when >50% scored 1 (green) and ≤15% 3 (orange), adjusted when >50% scored 2 (yellow) and ≤15% 1 (green), and omitted when >50% scored 3 (orange) and ≤15% 1 (green). If a score of an item fell outside these categories, the core group decided on retaining, adjusting or omitting the item. Number of golden tickets was scored per item. After this, in case of consensus across the focus groups (when all focus groups score the same), this score (retain, adjust, omit) was followed by the core group. If the scoring of an item was inconclusive, the core group decided on retaining, adjusting, omitting or merging items when meaning and content overlapped based on the memos qualitative input and taking the golden tickets into account. Items that were scored as ‘adjust’ were discussed by the core group and ultimately reformulated based on the written information on the memos and qualitative input. For the reformulation and merging of items the core group took enhancing readability and consistency of items throughout the questionnaire into account.

The core group (MS, JK, MB, CR, EB, JD) consisted out of six project members with diverse research backgrounds: a researcher with a background in psychology and statistics, a researcher with a background in population health, a researcher in the field of broad health concepts, a researcher in the field of health behavior, health promotion and public health, a researcher with a background in nutritional sciences and public health, and a researcher with a background in participatory research and shared decision-making. Step 1 led to a revised CPHQ with a reduced set of items.

### Step 2: Expert validation and member check

#### Expert validation

The revised CPHQ was validated with experts in two rounds: 1) with the core group together with experts involved in the research project 2) with experts from the national Positive Health research network covering a broad range of professionals. Finally, the core group discussed the input from these experts, and finally categorized the items into the components of the Capability Approach: ‘resources’, ‘conversion factors’, ‘functionings’ and ‘capabilities’ (Nussbaum, 2011) rendering a revised CPHQ version.

#### Member check

This second revised CPHQ version was submitted to 55 of the original 76 participants of the focus groups as only these had agreed on being contacted for further research. Participants were asked to give feedback per e-mail on the readability, comprehensibility and formulation of the items and headings using a 10-point scale (0 = worst, 10 = best). Furthermore, participants were asked their preference for one out of three answer options being 1) a 5-point Likert scale in words (checked for comprehensibility by the Dutch center of expertise in reducing health inequalities (PHAROS: www.pharos.nl)) or 2) a 5-point Likert-scale consisting of smiley icons or 3) a combination of both. Next, we asked for further suggestions. Participants were able to respond between 27-November-2023 en 19-December-2023. An overview of the member check questions is displayed in **appendix 2**.

## Results

### Step 1: focus group discussion results

**Table 1** shows characteristics of the participants of the focus groups. There was a tendency towards more professionals with work experience between 5-10 years, and more women who participated. Mostly highly educated people participated among professionals whereas among patients and citizens there were relatively more participants with a lower level of education.

**Table 1.**
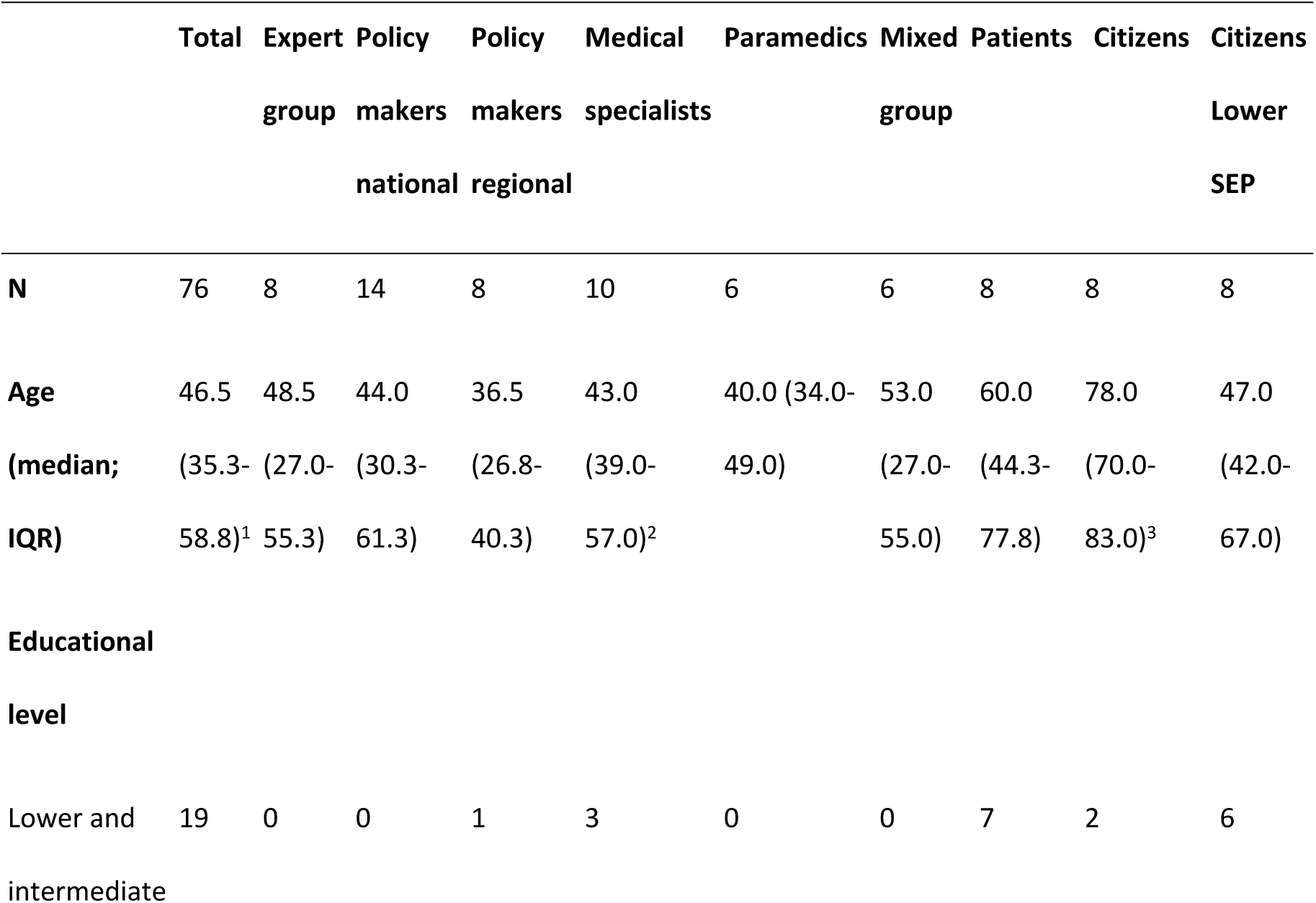
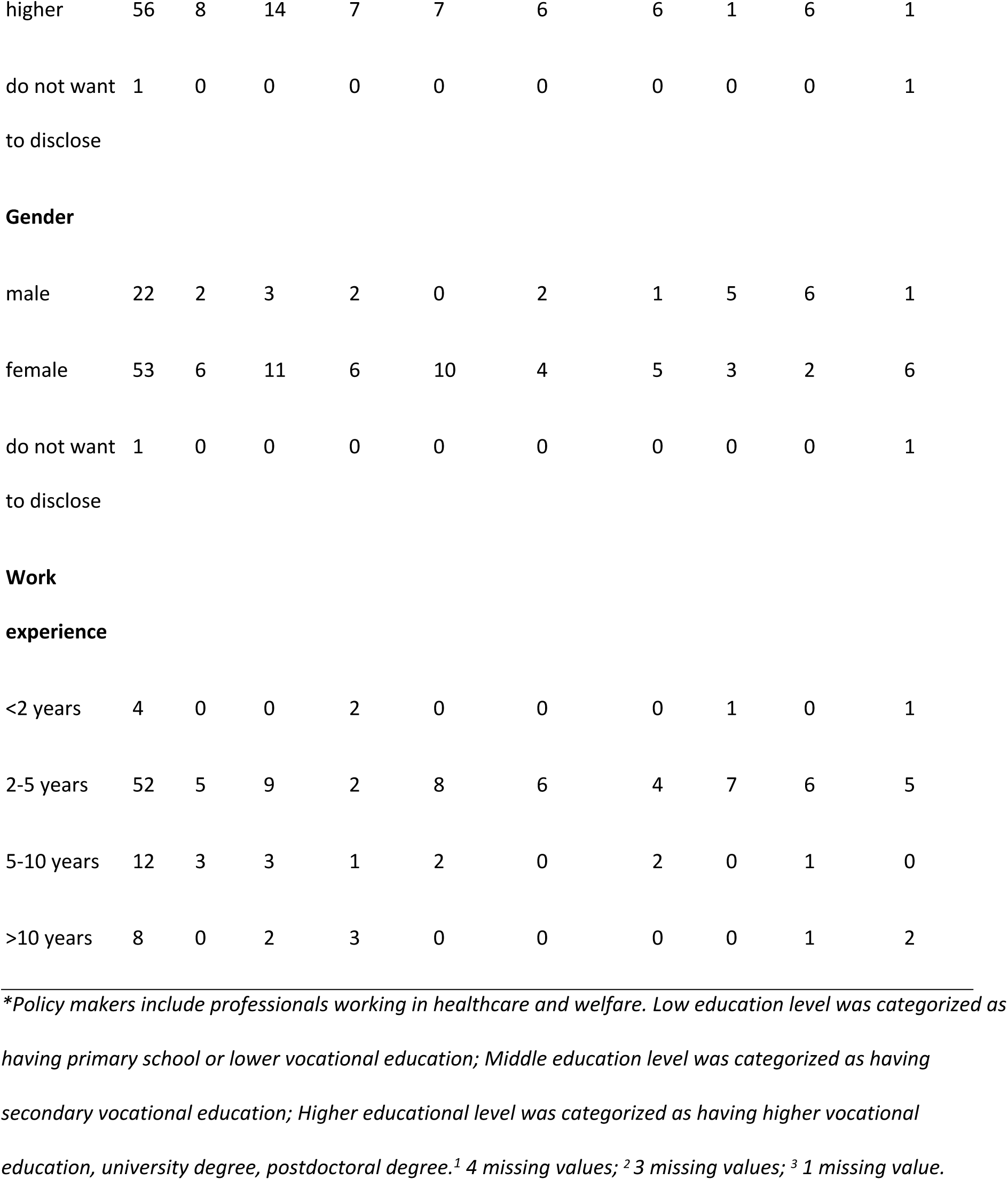
Overview and characteristics of the focus group discussion participants.

|  | Total | Expert group | Policy makers national | Policy makers regional | Medical specialists | Paramedics | Mixed group | Patients | Citizens | Citizens Lower SEP |
| --- | --- | --- | --- | --- | --- | --- | --- | --- | --- | --- |
| **N** | 76 | 8 | 14 | 8 | 10 | 6 | 6 | 8 | 8 | 8 |
| **Age** | 46.5 | 48.5 | 44.0 | 36.5 | 43.0 | 40.0 (34.0- | 53.0 | 60.0 | 78.0 | 47.0 |
| **(median;** | (35.3- | (27.0- | (30.3- | (26.8- | (39.0- | 49.0) | (27.0- | (44.3- | (70.0- | (42.0- |
| **IQR)** | 58.8)<sup>1</sup> | 55.3) | 61.3) | 40.3) | 57.0)<sup>2</sup> |  | 55.0) | 77.8) | 83.0)<sup>3</sup> | 67.0) |
| **Educational level** |  |  |  |  |  |  |  |  |  |  |
| Lower and intermediate | 19 | 0 | 0 | 1 | 3 | 0 | 0 | 7 | 2 | 6 |
| higher | 56 | 8 | 14 | 7 | 7 | 6 | 6 | 1 | 6 | 1 |
| do not want | 1 | 0 | 0 | 0 | 0 | 0 | 0 | 0 | 0 | 1 |
| Gender |  |  |  |  |  |  |  |  |  |  |
| male | 22 | 2 | 3 | 2 | 0 | 2 | 1 | 5 | 6 | 1 |
| female | 53 | 6 | 11 | 6 | 10 | 4 | 5 | 3 | 2 | 6 |
| do not want | 1 | 0 | 0 | 0 | 0 | 0 | 0 | 0 | 0 | 1 |
| experience |  |  |  |  |  |  |  |  |  |  |
| <2 years | 4 | 0 | 0 | 2 | 0 | 0 | 0 | 1 | 0 | 1 |
| 2-5 years | 52 | 5 | 9 | 2 | 8 | 6 | 4 | 7 | 6 | 5 |
| 5-10 years | 12 | 3 | 3 | 1 | 2 | 0 | 2 | 0 | 1 | 0 |
| >10 years | 8 | 0 | 2 | 3 | 0 | 0 | 0 | 0 | 1 | 2 |

*\*Policy makers include professionals working in healthcare and welfare. Low education level was categorized as having primary school or lower vocational education; Middle education level was categorized as having secondary vocational education; Higher educational level was categorized as having higher vocational education, university degree, postdoctoral degree.<sup>1</sup> 4 missing values; <sup>2</sup> 3 missing values; <sup>3</sup> 1 missing value.*

The qualitative analysis yielded input on several ‘overarching’ themes. These themes included “Formulation of items”, “Overlap between items”, “Redundant items”, “Importance of items”, “New suggestions”, and “Length/duration of the questionnaire”. **Table 2** shows an overview of the themes that emerged from the qualitative analysis and how this input was used for discussion within the core group and by experts for adjusting items after scoring the quantitative input.

**Table 2.** Themes that emerged from the focus group discussions about the original CPHQ.

| Theme | Explanation/examples |
| --- | --- |
| **Formulation of items** | Items in general, particularly related to health literacy, could be formulated shorter and easier to make them more understandable for people with limited literacy. For example the wording of some of the items that used “I am able to...” could be rephrased using alternative wordings such as “I can”.<br>-Examples or normative words such as “adequate” should be avoided.<br>-According to healthcare professionals items about feeling disadvantaged should be phrased positively. However, citizens and patients did not express this need and reported that they felt these items were important.<br>Uncertainty about how to interpret the word “environment” in items. It could indicate their street, neighbourhood, municipality but also their family and friends. This ambiguity made it difficult for the participants to answer these questions. They felt that the question should not be open for interpretation. |
| **Overlap between items** | Overlap was reported between;<br>- “It is hard for me to move on when something bad happens”, “I find it difficult to cope with stressful events”, and “It does not take me long to recover from a stressful event”<br>- “I can afford to live a healthy life” and “I can afford healthy food and exercise”<br>- items related to health literacy and access to health information: “I can communicate with healthcare workers and understand the explanation about my illness or treatment”, “It is clear to me where I need to go for medical help” and “When I look up or receive information about a subject, it is explained in a way that I can understand”. |
| **Redundant items** | - Political items should be avoided in a questionnaire to measure broad health |
| **Importance of items** | Most participants expressed that the items “I feel healthy”, “I feel fit”, and items related to relaxation were important items to measure health. |
| **New suggestions** | According to professionals an item related to a “sense of purpose” was missing. They suggested items that concerned ideals/goals in life or “I perceive my life as meaningful”.* However, citizens and patients did not express the need for such an item. |
| **Duration/number if items of the questionnaire** | Participants expressed that 32 items were too much. |

\* These suggestions also received seven golden tickets from participants.

After the quantitative analysis, the overall outcome was that 8 items were retained, 2 items were omitted, 22 items were adjusted of which 7 items were merged with other items. This led to a total of 23 items. See **table 3** for an overview of the quantitative results. In total, 76 golden tickets could be given of which 22 were given to “I feel healthy” and the others were distributed across diverse items without a clear preference. Furthermore, six participants did not use their golden ticket and 13 golden tickets were given to items from the ‘hand book’ or their own self defined items.

**Table 3.**
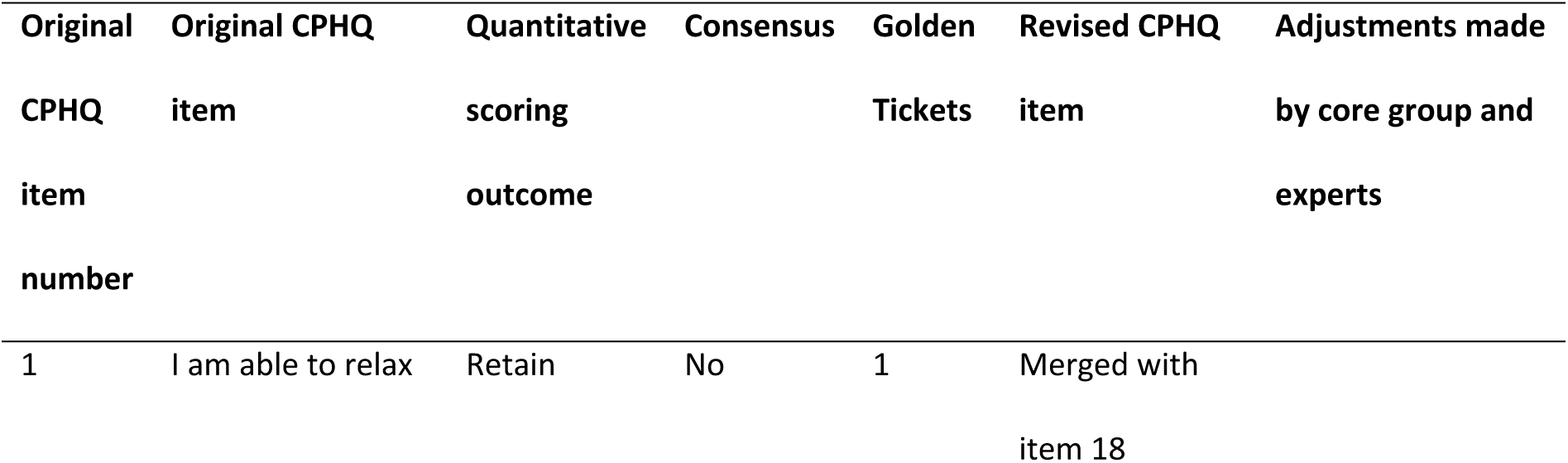
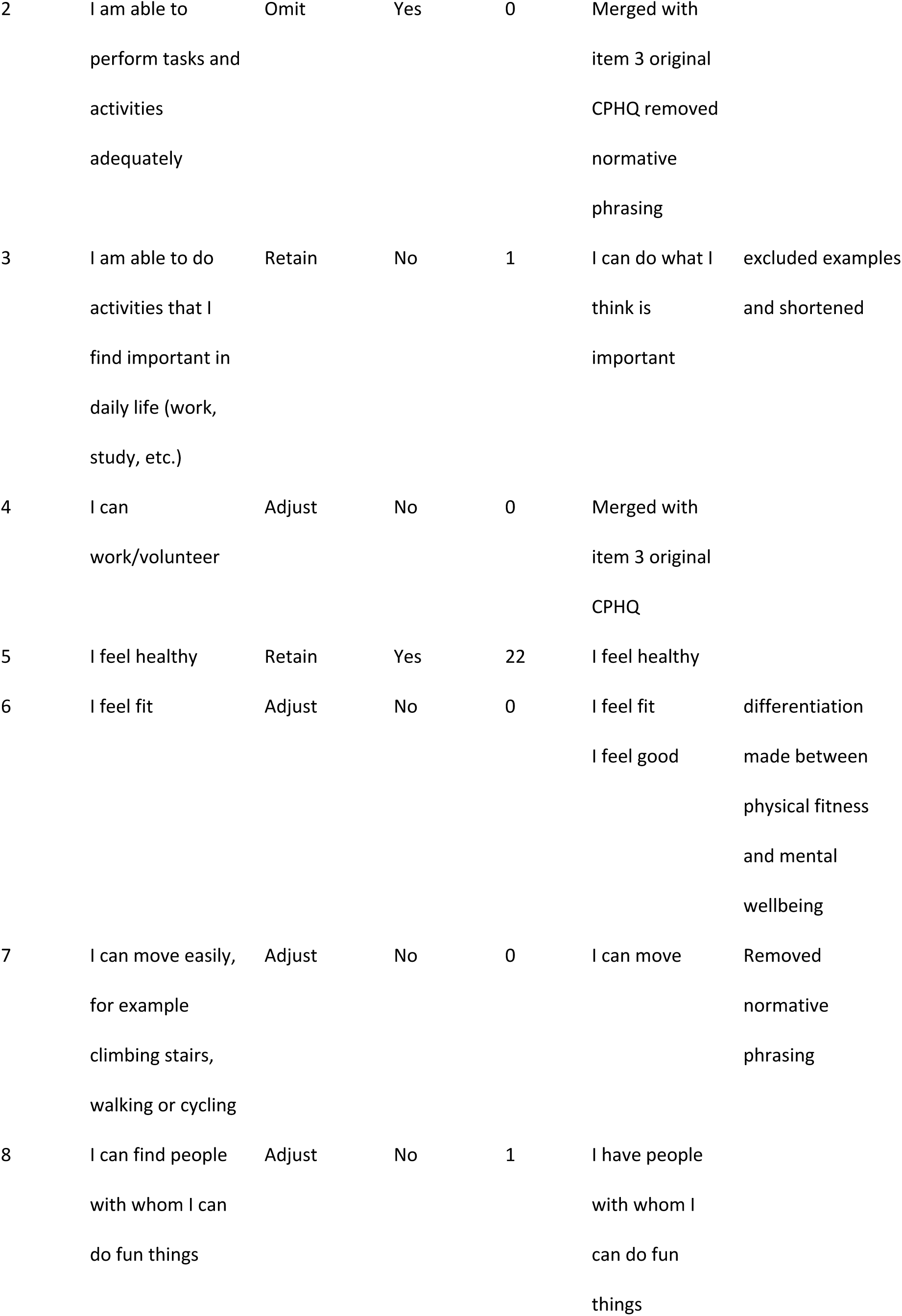
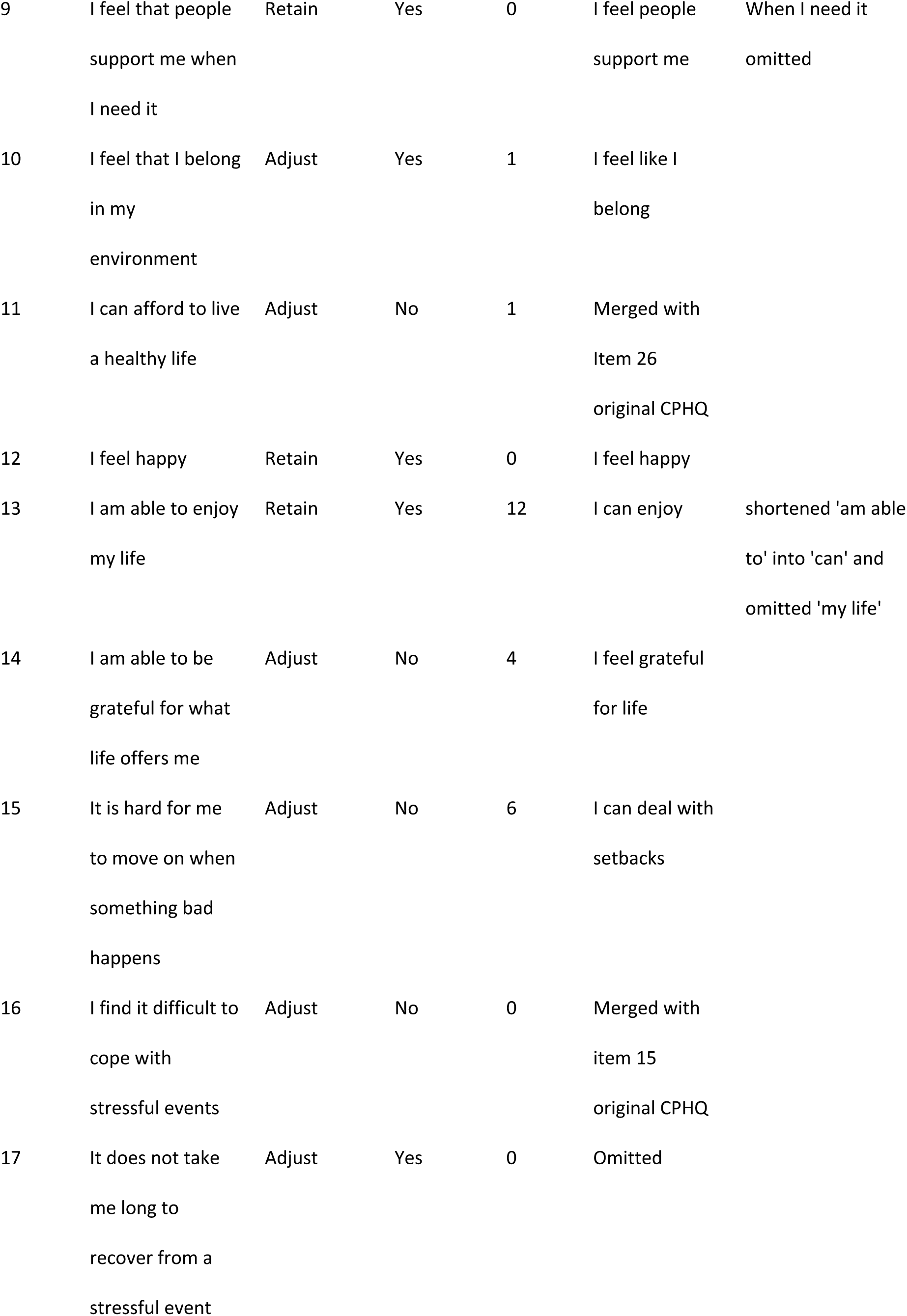
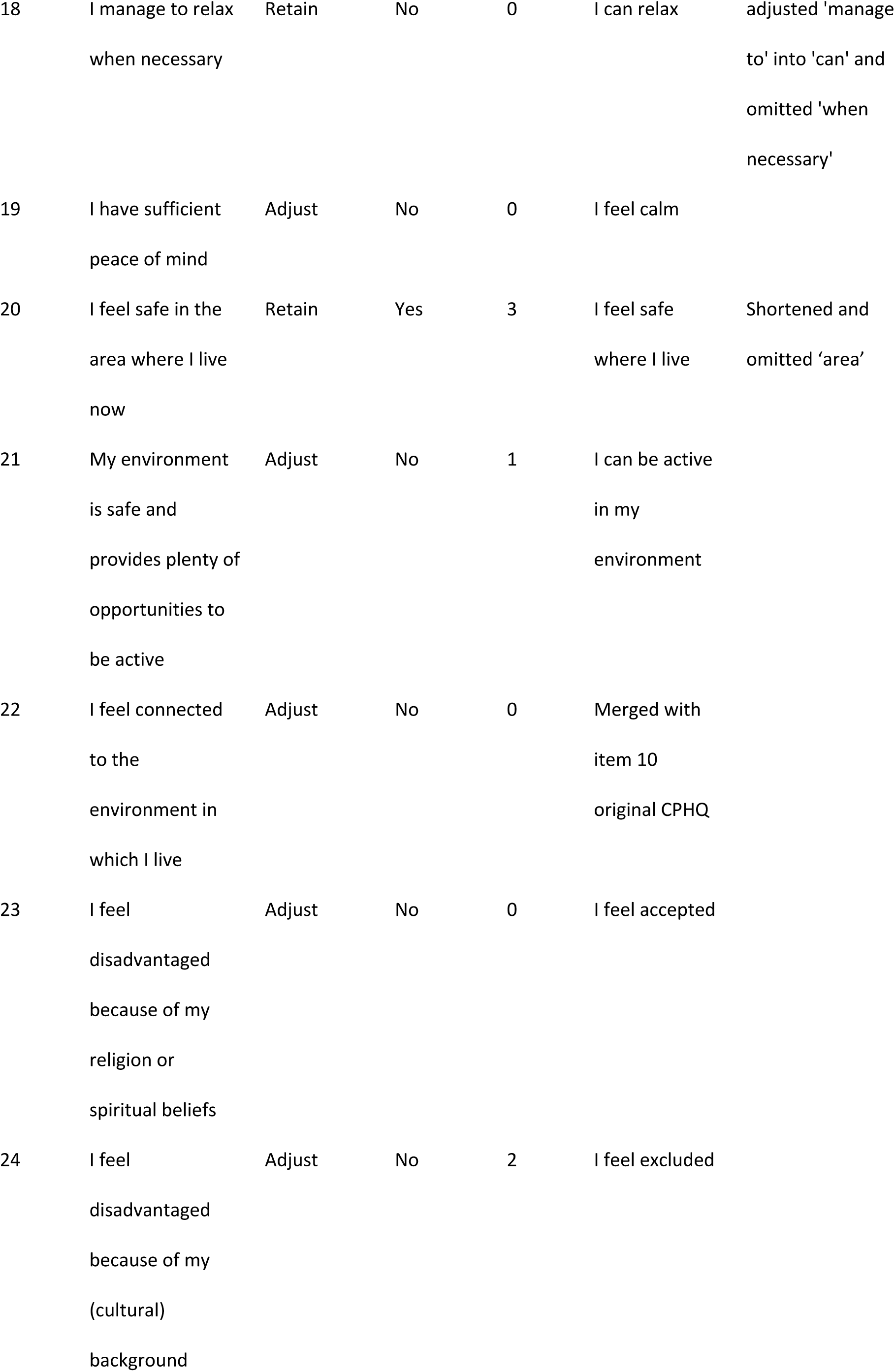
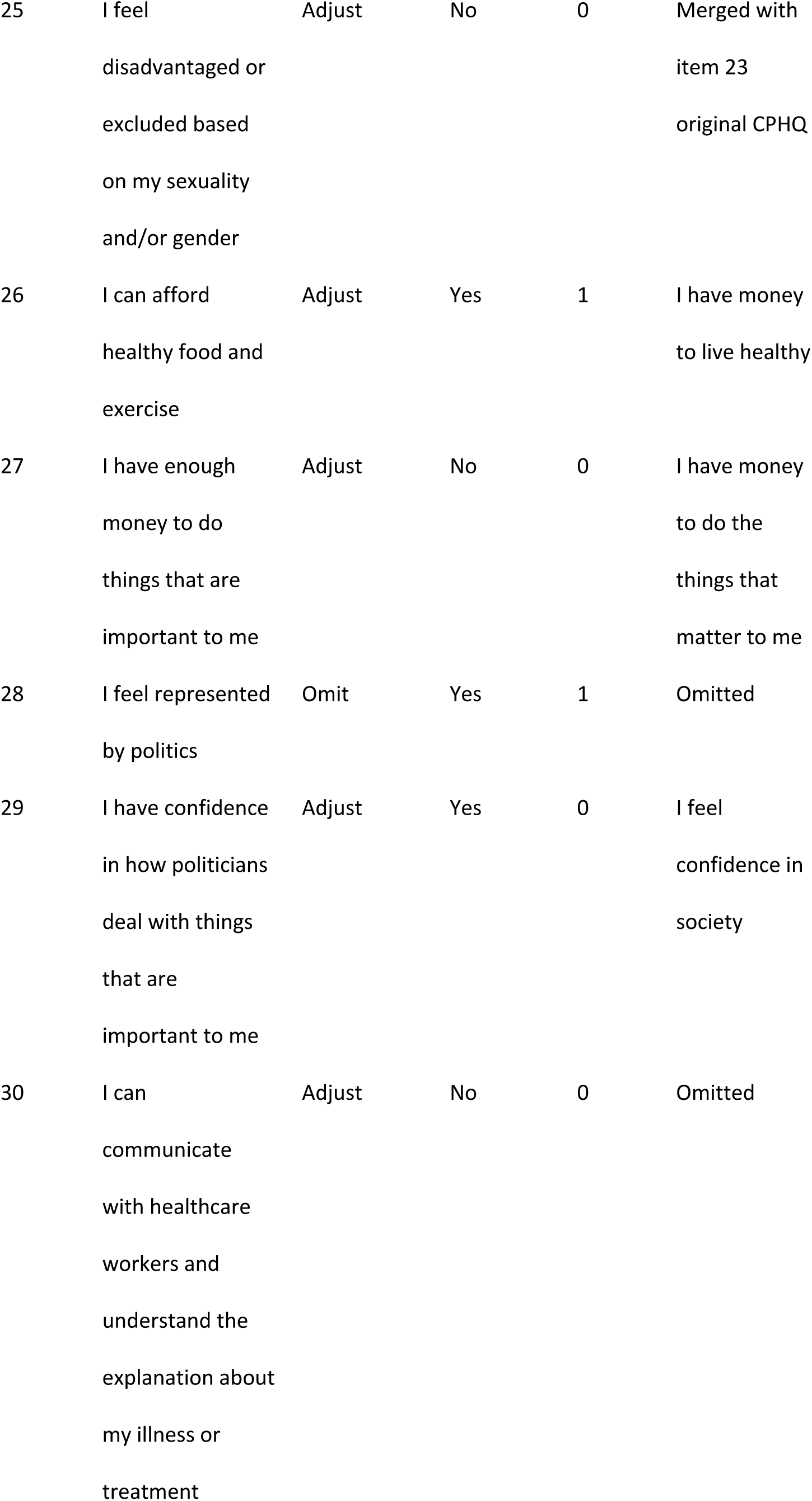
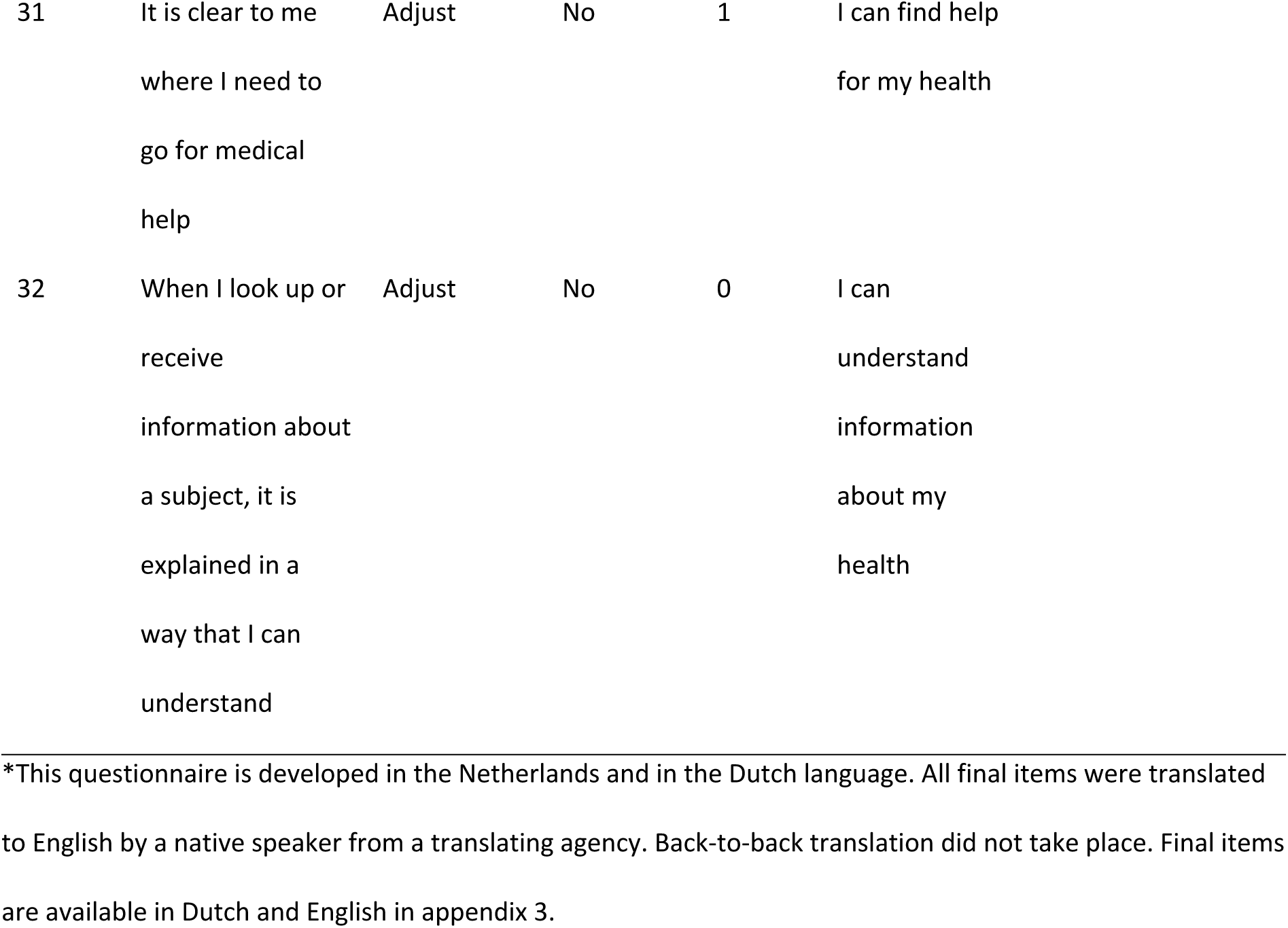
Quantitative results of the focus group discussions.

### Step 2: Expert validation and member check results

#### Expert validation results

The expert validation led to some refinement in formulation of items e.g. all phrased in a similar way. Based upon the expert validation the core group made a couple of final adjustments (See **table 3**). In line with the capability construct and the preferred short items, “I am able to enjoy my life” and “I manage to relax when necessary” were adjusted to “I can enjoy” and “I can relax”, respectively. Indicative words such as “my life” and “when necessary” were left out since enjoyment and relaxation are not dependent on those indications and can be filled in by personal circumstances. Similarly, for the sentences “I feel that people support me when I need it” and “I feel safe in the area where I live now”, the words “when I need it”, “area” and “now” were omitted. For the question “I am able to do activities that I find important in daily life (work, study, etc.)”, the core group decided to omit the examples and change the word activities in a more general verb “can”, formulating the new question as: “I can do what I think is important”. For the item “I feel fit”, there was a lot of debate whether this referred to physical fitness or mental fitness. Therefore, “I feel fit” was left in the questionnaire referring to physical fitness and the additional item “I feel good” was added, referring to mental wellbeing.

Furthermore, based on expert validation and discussion within the core group (with input of the focus group discussions) five items, that were previously omitted based on the COMET decision rules, were adapted and added doing justice to the broad concept of health. See **table 4** for an overview of the additional five items and reasons for adding these to the revised CPHQ. This increased the list of 23 items to 28 items.

**Table 4.**
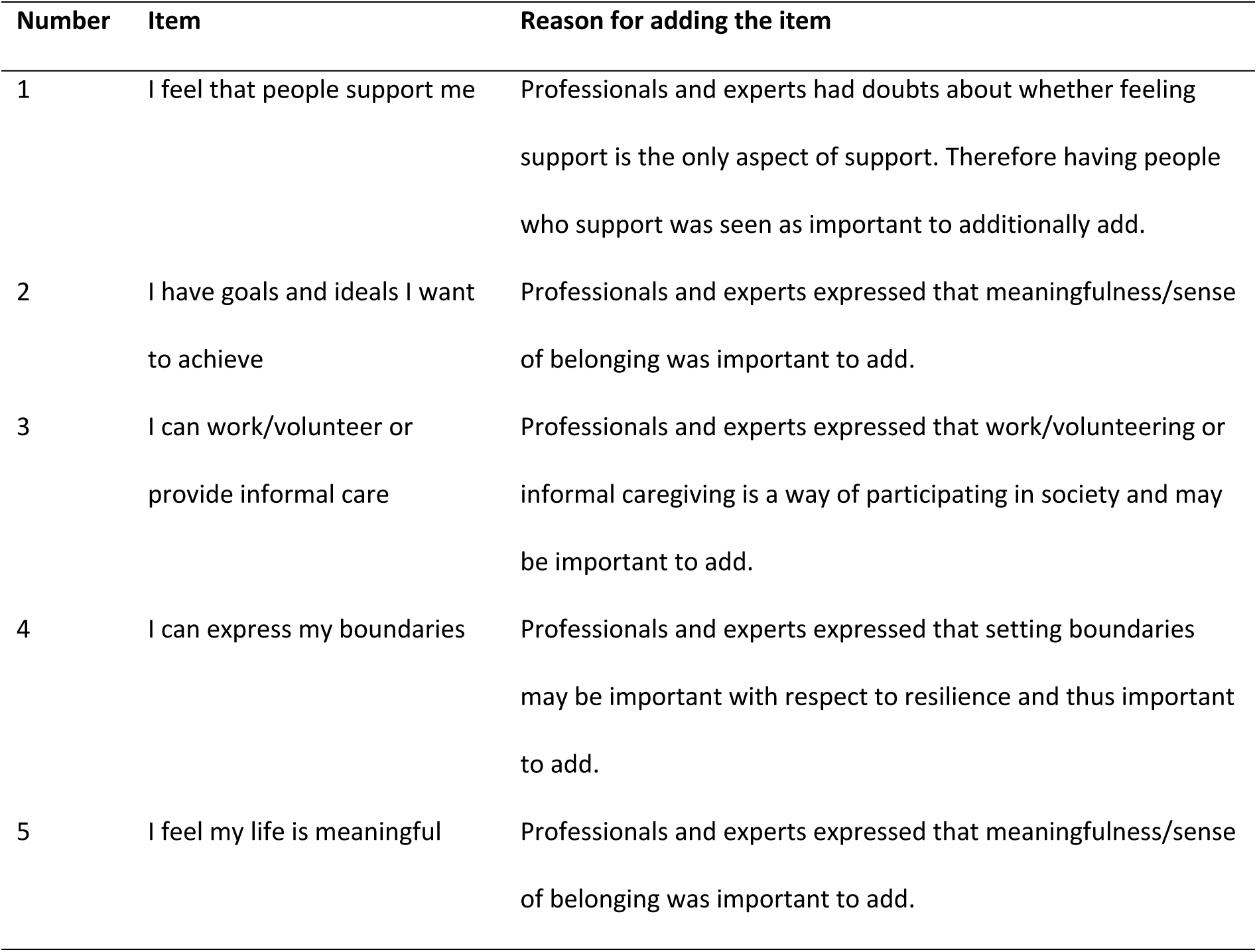
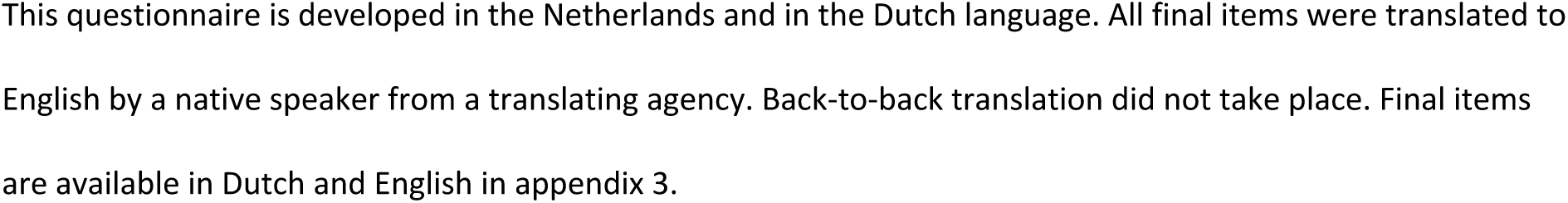
Additional items added after expert validation and core group discussion.

Subsequently, these 28 items were categorized into the components of the Capability Approach (**Table 5**) five items were categorized as ‘resources’ and phrased accordingly (e.g. “I have people […])”. Seven questions were categorized as conversion factors, referring to factors that influence how people can convert resources into capabilities. These were phrased as for example “I feel people support me”. Ten items were categorized as capabilities and phrased using “I can…”. Six items were categorized as functionings, related to their physical or mental state, and phrased as for example “I feel happy”. Categorization according to the Capability Approach was done because experts felt that the CPHQ questionnaire should be based on theory.

**Table 5.**
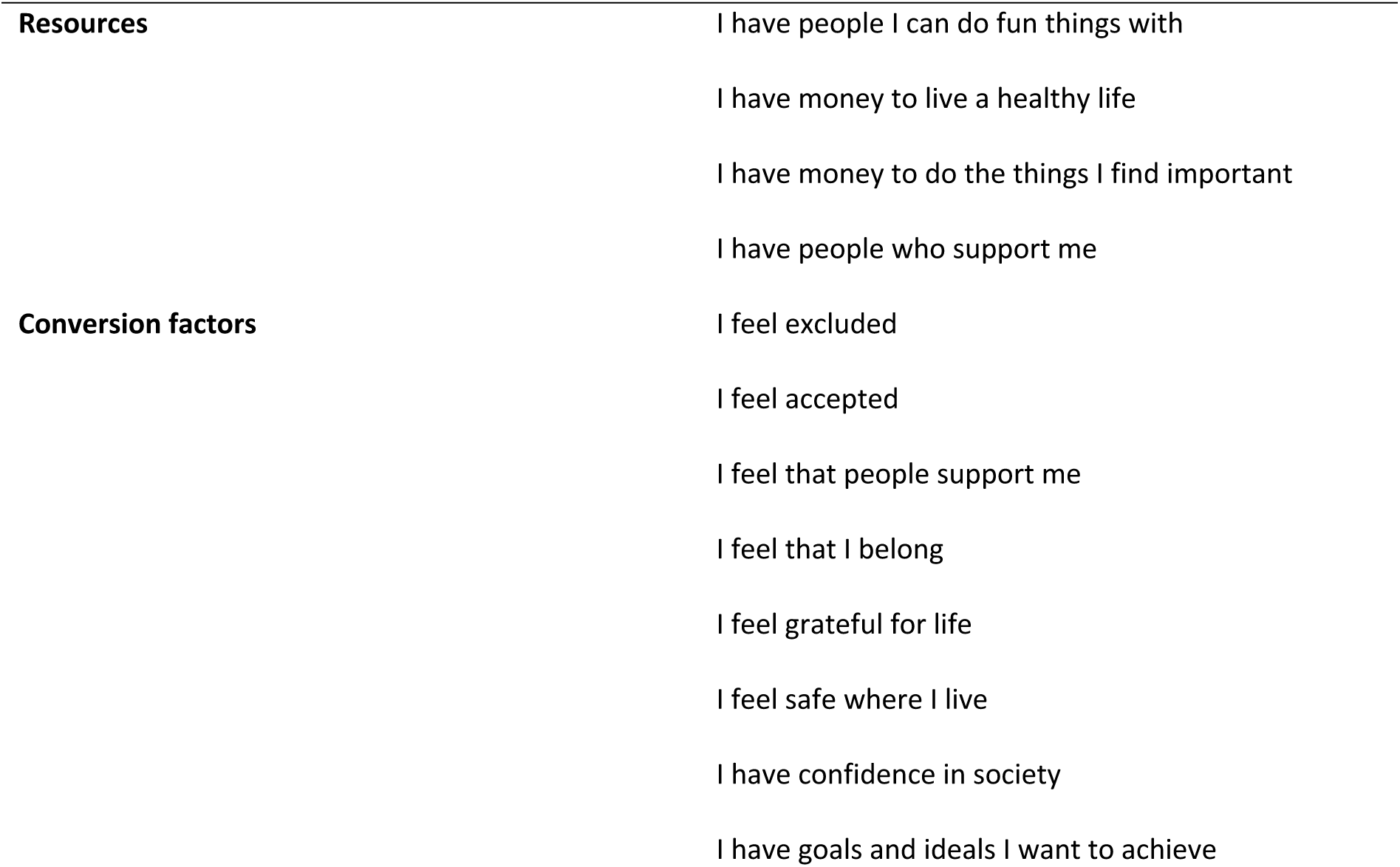
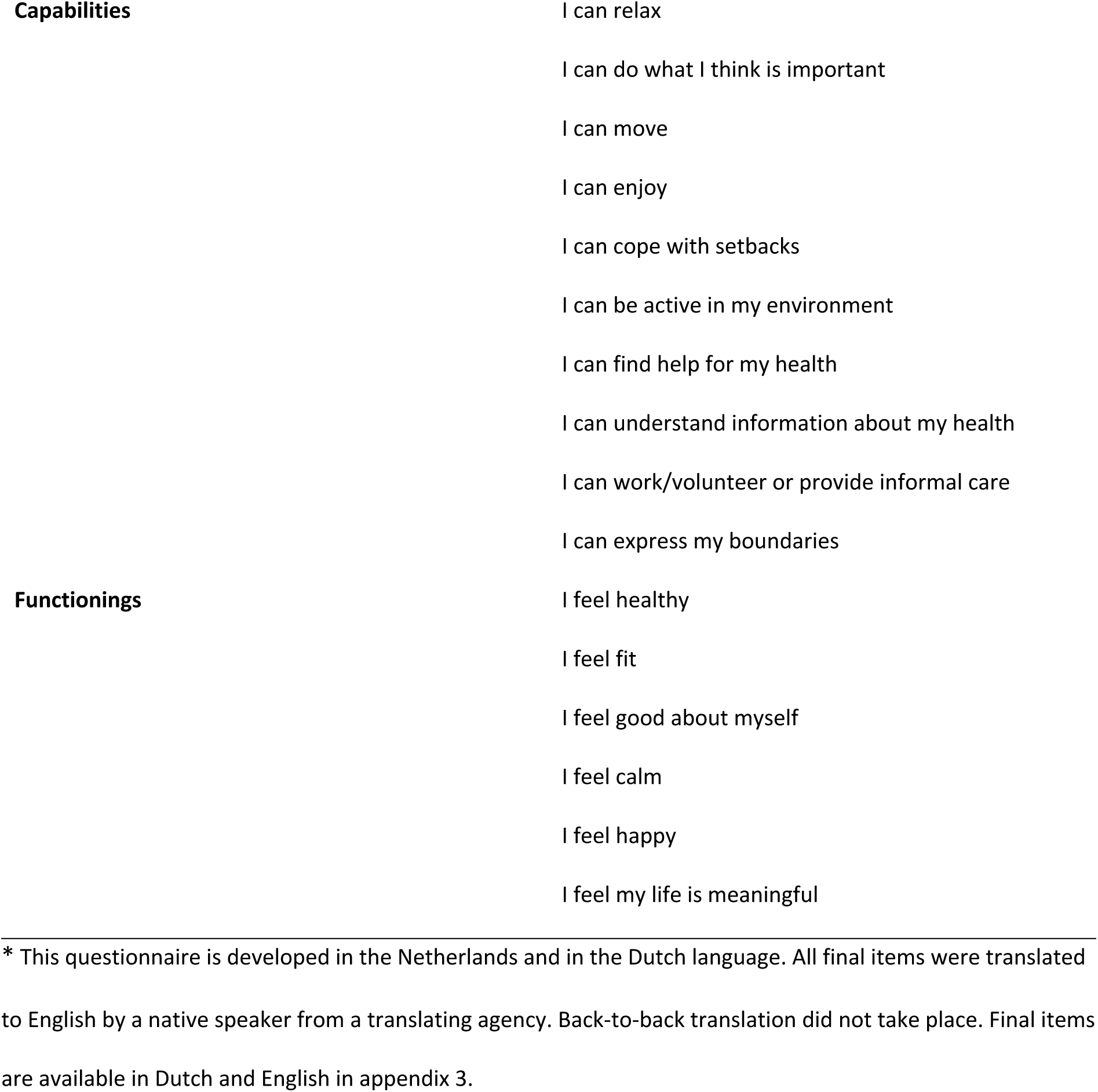
Items of the revised CPHQ categorized according to the Capability Approach.

As the questionnaire was developed in the Netherlands, the original items are in Dutch. These can be found in **Appendix 3**.

#### Member check results

The member check was sent to 55 of the 76 participants of the focus groups discussions participants who provided their email address and agreed to be contacted for additional information. The member check was conducted in the period between 27 November 2023 and 19 December 2023. In total, we received 18 replies (33%). Readability, comprehensibility and headings all scored good (see **table 6**).

**Table 6.** Results of the member check.

| Score of the questionnaire/items* | Mean±SD |
| --- | --- |
| Readability | 8.0±1.6 |
| Comprehensibility | 8.2±1.1 |
| Subheadings | 7.5±1.7 |
| **Preference of answer options\*\*** | Number (%) |
| Likert-scale in smileys | 8 (44%) |
| Likert-scale in text | 5 (28%) |
| Likert-scale with a combination of smileys and text | 2 (11%) |

\*Scored on a scale between 0-10 (i.e. not good to very good)
\*\*3 participants gave no answer on preferred answer option

There was no consensus based on what kind of answer options would be best suitable by the participants (see appendix 2). Given the various preferences, the core group decided for the most inclusive option being textual answers on a five-point Likert-scale accompanied with smileys on a five-point Likert-scale.

Suggestions of participants concerned 8 suggestions on the formulation, 3 on the grouping of items and 1 remark on the applicability for secondary care. With regard to the grouping and the applicability, no adjustments were made since the capability essentials were included in the grouping and the instrument is designed also for other domains than secondary care. With regard to the formulation, two participants questioned whether the opposite item “I feel excluded” would need to be stated in a positive formulation. Other remarks concerned the interpretation of words such as ‘grateful’ and ‘calm’, which is preference-sensitive. Given the fact that we did not receive any major concerns, we left the formulation unchanged.

## Discussion

This study aimed to develop a revised version of the 32-CPHQ to measure broad health. The resulting 28-item version is intended for use across multiple domains, including the healthcare and welfare policy domain, research and with specific attention to vulnerable populations (e.g. individuals with a lower SEP). The development process involved stakeholders from all aforementioned domains and builds on Positive Health and the Capability Approach. This ensures that the revised-28 item CPHQ is theoretically grounded, reflects stakeholder and end-user perspectives and ensures content validity (Ricci et al., 2019)

### Key findings

#### Commonalities

One key point across all focus group discussion was the consensus to avoid examples and normative phrasing in questionnaire items. This allows for more subjective interpretation by the respondents. For instance, the item “I am able to do activities that I find important in daily life (work, study, etc.)” originally included examples, but participants emphasized that what is considered important varies across individuals and contexts (Robeyns, 2005, 2013). Allowing room for personal interpretation aligns with the aim to measure health in a context-sensitive way. However, it may result in their answers reflecting different underlying constructs meaning that the item can no longer reliably measure a single, clearly defined concept. If so, this reduces the internal consistency. Further research is needed to examine this.

Another commonality was that the item “I feel healthy” was perceived as the most important item by participants. Other frequently emphasized items included “I feel fit,” “I feel good,” and “I feel happy”. Interestingly, these items reflect functionings in the terms of the Capability approach: they describe actual states of being and doing (Nussbaum, 2011). Some of these items such as “feel healthy” or “I feel happy” are also used as standalone indicators in instruments that measure health, quality of life or well-being. For instance, the EQ-5D-5L includes a visual analogue scale (VAS) where respondents rate how healthy they feel, and single-item measures or happiness are also used in population health surveys (Andrews & Withey, 2012; Feng et al., 2021). This suggests that such functionings may not only serve as outcomes but also as overarching, integrative indicators of health. However, according to the Capability Approach, what matters is not only the realization of these experienced states but the freedom and opportunity (i.e. capabilities) to achieve them. Instruments that focus solely on functionings may miss whether people actually had the opportunity to achieve these states, while instruments that only assess capabilities might miss how people actually perceive their health. Therefore, we suggest that to assess broad health, a measurement instrument should include both elements.

Participants also discussed the item “I feel represented by politics”, and were in agreement that it should be removed from the questionnaire-but for different reasons. Medical professionals questioned its relevance, as it lies beyond their professional influence. Patients and citizens felt that it was unrelated to their personal health. Interestingly, a related item on trust in society remained in the revised CPHQ. Previous research shows a link between poor health with lower political trust (Mattila, 2020). Mattilla (2020) suggests that one reason for this lower trust might be a disconnect between what citizens expect from the government and institutions and how they actually perceive its outcomes, particularly regarding how fairly they feel treated (Mattila, 2020). From a Capability Approach perspective, political representation can act as a conversion factor that shapes people’s ability to achieve valued states (Robeyns, 2005). Moreover, in the original CPHQ, items on political representation formed a distinct factor in the factor analysis (Doornenbal et al., 2024). These differing perspectives reflect the complexity of determining which contextual and societal factors should be included in a measure of broad health. We therefore recommend that future validation studies further explore whether and how aspects like political representation and trust in institutions contribute to people’s perceived capabilities for (broad) health. Rather than including these items by default, their relevance may need to be assessed contextually, depending on the population or on the purpose of the measurement.

#### Differences

There were also some key differences between the stakeholder groups. End-user respondents (i.e. (lower SEP) citizens and patients) preferred the item “I feel excluded” to be phrased negatively as they did not recognize themselves in its positive form. This reflects findings from the original CPHQ development(Doornenbal et al., 2024). Professionals, researchers and experts, however, favored positive phrasing. This divergence may reflect differences in how health is conceptualized across social strata. Several studies suggest that individuals with lower education levels are more likely to define health in negative terms (Dierx & Kasper, 2022; Peersman et al., 2012; Platzer et al., 2021; Stronks et al., 2018). To illustrate, Peersman et al. found that lower-educated respondents referenced the presence of health problems, while higher-educated individuals emphasized the absence of such problems. The differences may stem from both cultural framing and real differences in health experiences across socioeconomic groups (Stronks et al., 2018). To ensure that health measurement tools are inclusive and sensitive to different lived realities it may be useful to examine how individuals from different SEP strata interpret specific items, particularly those related to exclusion or disadvantage. While we already included citizens and low SEP citizens, future development and implementation efforts may benefit from continued attention to subgroup differences in interpretation of item phrasing.

Another difference concerned the inclusion of the concept of meaningfulness. Where professionals, researchers and experts emphasized its importance, citizens and patients did not mention meaningfulness. This may be due to the abstract nature of the concept, which may not surface spontaneously in discussions. Nonetheless, meaningfulness has been emphasized in the literature on Positive Health, even by patients (Huber et al., 2011; Huber et al., 2016). Also in earlier research, in the concept of salutogenesis, which is about the origins of health and focuses on factors that support health and well-being, developed by Antonovsky, meaningfulness is seen as an important asset for health (Moksnes, 2021). Following expert consultation and discussion within the core group, an item on meaningfulness was added to the revised questionnaire. Future psychometric analysis will determine whether it should be retained.

### Strengths and limitations

This study has several strengths. First, the questionnaire was developed through co-creation with diverse stakeholders from healthcare, welfare, and policy domains and including patients and (lower SEP) end-users. This user-centered approach increases the likelihood of acceptance and applicability in various settings. Second, the inclusion of participants with a lower SEP and from different regions of the Netherlands improves the relevance and representativeness of the instrument. Third, the iterative development process including expert validation and a member check enhanced the credibility and robustness of the 28-item CPHQ. Fourth, the questionnaire is theoretically grounded not only in Positive Health but also in the Capability Approach, which adds more attention to contextual influences on health. Fifth and finally, recurring themes across focus group discussions suggest that thematic saturation has been reached.

Nevertheless, some limitations should be acknowledged. There may be some recruitment bias towards individuals who are already more positive towards the concept of broad health. Additionally, consensus was not always reached for specific items. Some decisions with respect to wording of items were therefore compromises and may not be ideal formulations for all stakeholders. However, a member check was performed for contingency.

### Implications and future research

The revised CPHQ offers a promising tool to measure broad health. It could provide valuable insights in the effectiveness of health and welfare interventions or (policy) programs that are currently assessed from a more narrow perspective. Furthermore, such a measurement instrument may be used for population monitoring. However, it is a challenge to measure broad health with a compact questionnaire. In further research we will investigate if the revised CPHQ is sufficiently sensitive to pick up small changes in health status.

Next steps in the validation of the revised CPHQ include item reduction analysis (e.g. by factor analysis) to assess the most important items and to potentially shorten the questionnaire. Questionnaire length can have a serious impact in how the questionnaire is filled out and on response rates (McColl et al., 2002). Discriminant validity should be assessed by examining scores on the revised CPHQ across different subpopulations. Examining correlations with other health questionnaires (e.g. EQ-5D-5L) and analyzing changes in scores over time will further support its validity. Furthermore, future research should focus on its responsiveness in diverse populations and settings. This is in line with a recent rapid review on the overall concept of Positive Health which emphasizes the need for more clarity regarding suitable settings and target groups for the implementation of Positive Health, with special attention paid to the inclusion of vulnerable populations (van Vliet et al., 2024). Once the revised CPHQ has been evaluated and proven useful in several subpopulations and settings, it would be interesting to investigate how the revised CPHQ relates to other questionnaires measuring broad concepts of health such as the 17-item Positive Health questionnaire and whether it is possible to adapt it to other populations such as youth.

## Conclusions

In conclusion, the revised CPHQ was developed with input from a wide range of stakeholders including patients and (lower SEP) citizens. This led to a 28-item questionnaire to measure broad health. Future research, through cross-sectional or longitudinal research in varying populations and settings has to ensure its validity.

## Data Availability

Data cannot be shared publicly because of privacy. Data are available from the Leiden University Medical Center Institutional Data Access (contact via the corresponding author) for researchers who meet the criteria for access to confidential data.

## Acknowledgments

The authors are grateful for the participation of all participants to the focus groups and the experts from the national Positive Health research network for providing constructive feedback. The authors are especially grateful for the contribution of research assistant Ine Hesdahl-de Jong for assisting in the focus groups and structuring the data.

